# Characterizing superspreading of SARS-CoV-2 : from mechanism to measurement

**DOI:** 10.1101/2020.12.08.20246082

**Authors:** Zachary Susswein, Shweta Bansal

## Abstract

Superspreading is a ubiquitous feature of SARS-CoV-2 transmission dynamics, with a few primary infectors leading to a large proportion of secondary infections. Despite the superspreading events observed in previous coronavirus outbreaks, the mechanisms behind the phenomenon are still poorly understood. Here, we show that superspreading is largely driven by heterogeneity in contact behavior rather than heterogeneity in susceptibility or infectivity caused by biological factors. We find that highly heterogeneous contact behavior is required to produce the extreme superspreading estimated from recent COVID-19 outbreaks. However, we show that superspreading estimates are noisy and subject to biases in data collection and public health capacity, potentially leading to an overestimation of superspreading. These results suggest that superspreading for COVID-19 is substantial, but less than previously estimated. Our findings highlight the complexity inherent to quantitative measurement of epidemic dynamics and the necessity of robust theory to guide public health intervention.

## Introduction

Heterogeneity in individual transmission is a critical feature of infectious disease dynamics, enabling explosive superspreading events. Although superspreading was identified during the HIV pandemic [1], it is since the SARS-CoV-1 outbreak in 2002-2003 that superspreading has become a central focus of investigation; its large transmission clusters were not well-explained by traditional epidemic theory [2, 3] and the public health response to curtail spread required a novel approach [4, 5]. In the twenty years after the SARS-CoV-1 outbreak, advances in theory have enabled quantitative description of superspreading potential, but disentangling the behavioral and biological mechanisms driving superspreading remains elusive and little attention has been paid to potential biases in superspreading measurement. Superspreading has been identified as key to SARS-CoV-2 transmission and understanding and accurately measuring superspreading are crucial for an informed public health response.

A key insight in the theory of superspreading has been the definition of the offspring distribution as a negative binomial random variable [6]. The offspring distribution, describing the probability that an infected case will transmit to a given number of secondary cases, is quantified by a mean, *R*_0_, and a dispersion statistic, *k*. Accurate measurement of the offspring distribution enables quantitative description of the rate and manner of a pathogen’s spread. High values of the offspring distribution’s dispersion statistic (*k >* 5) suggest relatively homogeneous transmission. On the other hand, low values of *k* describe high variability in transmission, with most primary infections leading to few new cases and a few rare primary cases producing many secondary cases. This dispersion statistic has become widely used as a measure of superspreading potential, with values of *k* less than one indicating substantial superspreading [7].

Accurate computation and measurement of this dispersion statistic provides critical insight into the nature of pathogen transmission, informing actions as disparate as predictive modeling and public health control measures [7, 8]. Offspring distributions well-explained by low values of *k* correspond to the 80-20 rule, which posits that 80% of new infections are attributable to 20% of individuals [9, 10]. Proposed control measures take advantage of this heterogeneity by preferentially targeting these individuals, for example by preventing public congregations and thereby preventing superspreading events [10, 9]. Estimation of *k* (and empirical adherence to the 80-20 rule) can also guide resource allocation, with low values of *k* suggesting greater return on investment from contact tracing – limiting transmission by preventing superspreading events before they occur [11]. Models and simulation studies endorse the utility of targeting this high-transmission group, but emphasize the challenge presented by pre-symptomatic transmission; isolation of symptomatic individuals and robust contact tracing may not be enough to control an outbreak if substantial transmission occurs before cases can be identified [12].

Superspreading is a function of heterogeneity in behavior and transmissibility [6, 13]. Human behavior can be variable across a population which can create disproportionate skews in disease transmission [14, 15, 16]. Similarly, between-individual variability in transmissibility, due to differences in susceptibility or shedding, can contribute to superspreading [17, 3]. Here, we encapsulate these many sources of heterogeneity into two factors, behavior and biology. Behavior is driven by contact between individuals, with heterogeneity arising from variation in age, occupation, social practices, environment, and the interaction type necessary for transmission, and leads to differences in numbers of contacts. We use “biology” to refer to differences in individual transmission competence (i.e. transmissibility), with heterogeneity arising from variation in human physiology (e.g. shedding or susceptibility), viral competence, and contact environment, among other factors. While these mechanisms contribute in complex ways to disease transmission dynamics, the relative contributions of behavior and biology to superspreading behavior are still largely unexamined, which limits the targeting and implementation of control measures.

During the SARS-CoV-2 pandemic there has been substantial interest in accurate measurement of the *k* statistic, quantifying the pathogen’s superspreading capability [18, 19, 20, 15]. Large clusters of infections have been identified as a key feature of SARS-CoV-2 transmission, characteristic of superspreading behavior [21, 22, 23, 19, 24]. Matching these reports, many empirical estimates find *k* to be quite low (∼ 0.1-0.4), suggesting substantial heterogeneity in the SARS-CoV-2 offspring distribution [19, 20, 25, 15]. Proposed control strategies take this heterogeneity into account, allowing for a potentially more efficient public health response [26, 13, 20, 27].

Here, we use a simple mechanistic model with behavior and transmissibility parameterized at the individual level to understand how heterogeneity in transmission is affected by variation in behavior versus transmissibility. Our results suggest that existing theory on the behavior that underlies the transmission of respiratory infections, like SARS-CoV-2, cannot explain many empirical measurements of superspreading potential (i.e. the *k* statistic). However, we show that methodological limitations and sampling errors could reduce the measured value of the *k* statistic, resolving this discrepancy. We apply these results to SARS-CoV-2 transmission, re-examining several publicly available contact tracing datasets to estimate the superspreading potential of COVID-19, and demonstrate that superspreading occurs substantially, but at a lower rate than many empirical estimates (i.e. higher *k*), consistent with theoretical results.

## Results

### Superspreading is driven by contact heterogeneity

To clarify the independent role of behavior versus biology in superspreading, we consider how heterogeneity in either factor impacts the *k* statistic, while controlling *R*_0_. As expected, variation in contact structure (as measured by the degree distribution) or variation in shedding or susceptibility (as measured by the transmissibility) generates increased variation in transmission (as measured by the *k* statistic of the offspring distribution) (Figure 1A). When there is heterogeneity in behavior alone, superspreading is observed but still not pronounced (*k* ≥ 1), with no observations at the extreme end of the empirically observed range (*k <* 0.5). Furthermore, equivalent levels of heterogeneity in transmissibility alone are not sufficient to produce superspreading (*k >* 2). (See Appendix Figure S1 for univariate sensitivity analysis).

**Figure 1:**
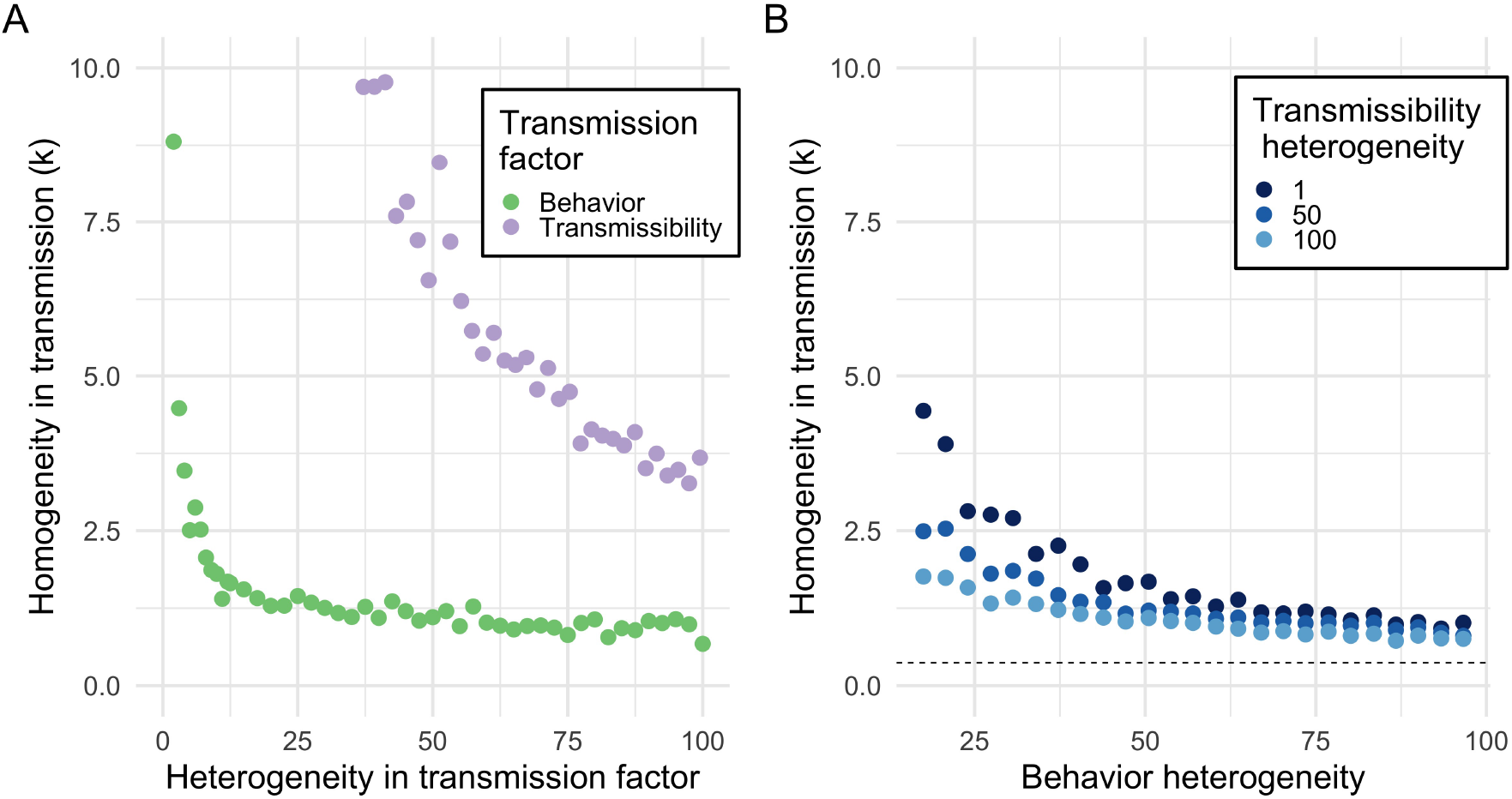
Superspreading is governed more by behavior (degree variance) than transmissibility a) Heterogeneity in behavior (green) or transmissibility (purple) alone produces moderate amounts of superspreading. Variation in contact structure alone (i.e. behavior) produces substantially more offspring dispersion than variation in transmissibility alone. However, neither process alone can account for *k <* 1. b) When combined, heterogeneity in behavior and biology can produce superspreading in the range 0.5 ≤ *k* ≤ 1, more than either process can alone. This superspreading behavior is largely governed by social contact patterns, with transmissibility only weakly modifying *k*. A power law degree distribution (extreme contact variation; dashed line) is needed to produce *k <* 0.5.

When we consider the more realistic scenario of simultaneous heterogeneity in the degree distribution and transmissibility (and allowing *R*_0_ to vary), we find that variation in the offspring distribution is largely governed by variation in contact rather than in transmissibility (Figure 1B). This relationship is only weakly sensitive to the mean transmissibility or degree, while other epidemic statistics, like the *R*_0_, are more sensitive to the means. The synergy of heterogeneity produced by both factors can result in moderate superspreading (0.5 ≤ *k* ≤ 1) beyond what either factor produces separately, but only extreme contact heterogeneity (a scale-free degree distribution) achieves extreme superspreading (*k* ≤ 0.5). (See Appendix S2 and S3 for bivariate sensitivity analysis).

Our analysis also produces a functional relationship between the *k* statistic and the 80%/20% rule (Figure 2), and we find that values of *k* between 0.5 and 1 largely underestimate this proportion, instead leading to 60-70% of secondary infections caused by 20% of primary cases. Indeed, we find that the 80%/20% rule is satisfied only with extreme variability in the offspring distribution (*k* ≈ 0.3). Past empirical results have found 80%/20% rule adherence across a range of 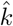 estimates, both higher and lower than the theoretical relationships we demonstrate. We hypothesize that this discrepancy may be explained by variability in measurement and selection bias, and investigate this below.

**Figure 2:**
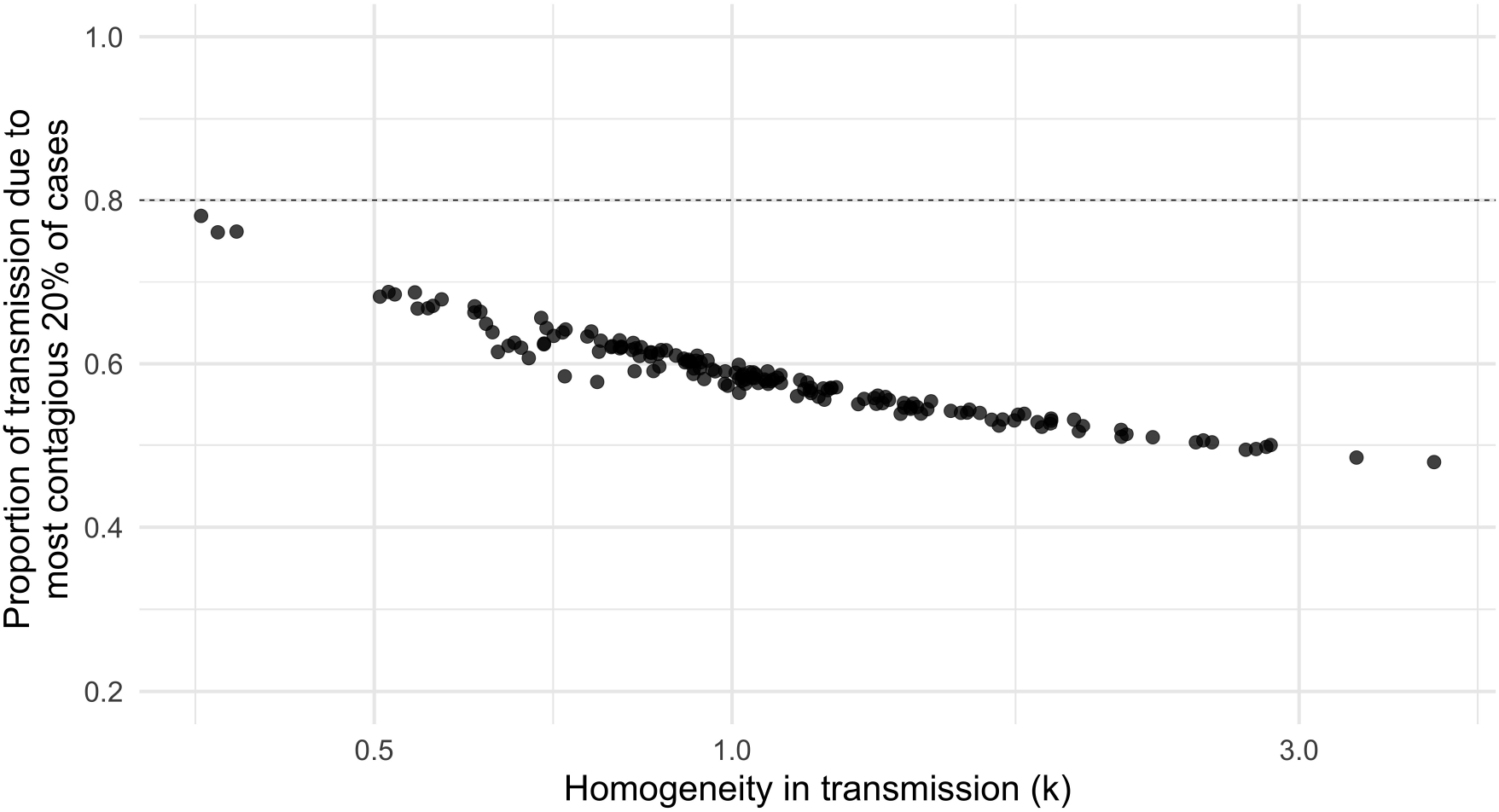
A *k* value of 0.3 corresponds to the 80/20 rule. Simultaneous heterogeneity in behavior and transmissibility produce *k* values that correspond to substantial variation in per-individual rates of transmission. Values of *k* close to 1 correspond to 50% of secondary cases caused by 20% of primary cases. Values of *k* in the range of 0.3 are consistent with 80% of secondary cases caused by 20% of primary cases – adherence to the 80/20 rule. Dashed line indicates 80/20 rule adherence.

### Superspreading measurement is noisy and biased

Quantitative estimation of superspreading is methodologically challenging. We find that even with true random samples at reasonably large sample sizes (200 primary cases, corresponding to roughly 500-1000 observed secondary cases), the point estimate of the *k* statistic (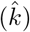) is often imprecise, with approximately 50% of estimates not within 10% of the true mean (Figure 3A). Additionally, we find that 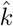 is equally dispersed around the true population parameter, with standard error increasing with the magnitude of the true population *k* (see Appendix Figure S4). Therefore, larger population values of *k* can often produce small values of 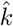, leading to overestimates of superspreading purely due to sampling variation. While confidence intervals can ameliorate issues presented by noisy point estimates, the quality of confidence interval coverage guarantees depends strongly on the method of estimation. In Figure 3B, we show that Wald confidence intervals (which assume convergence to normality) perform poorly when the true *k* value is small – the scenario of interest for superspreading pathogens. Larger sample sizes improve the convergence rate, but when the true value of *k* is below 0.5 the coverage probability is still well below the nominal level. On the other hand, the non-parametric bootstrap method performs well across all the scenarios, with coverage closely approximating the nominal level. These results rely on the assumption of random sampling; 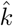 can be biased downwards with respect to the true *k* (i.e. overestimate superspreading) if datasets demonstrating superspreading behavior are selectively used to quantify *k*, a bias magnified by imperfect case observation (like the data produced by contact tracing; Appendix Figure S5, Table S1).

**Figure 3:**
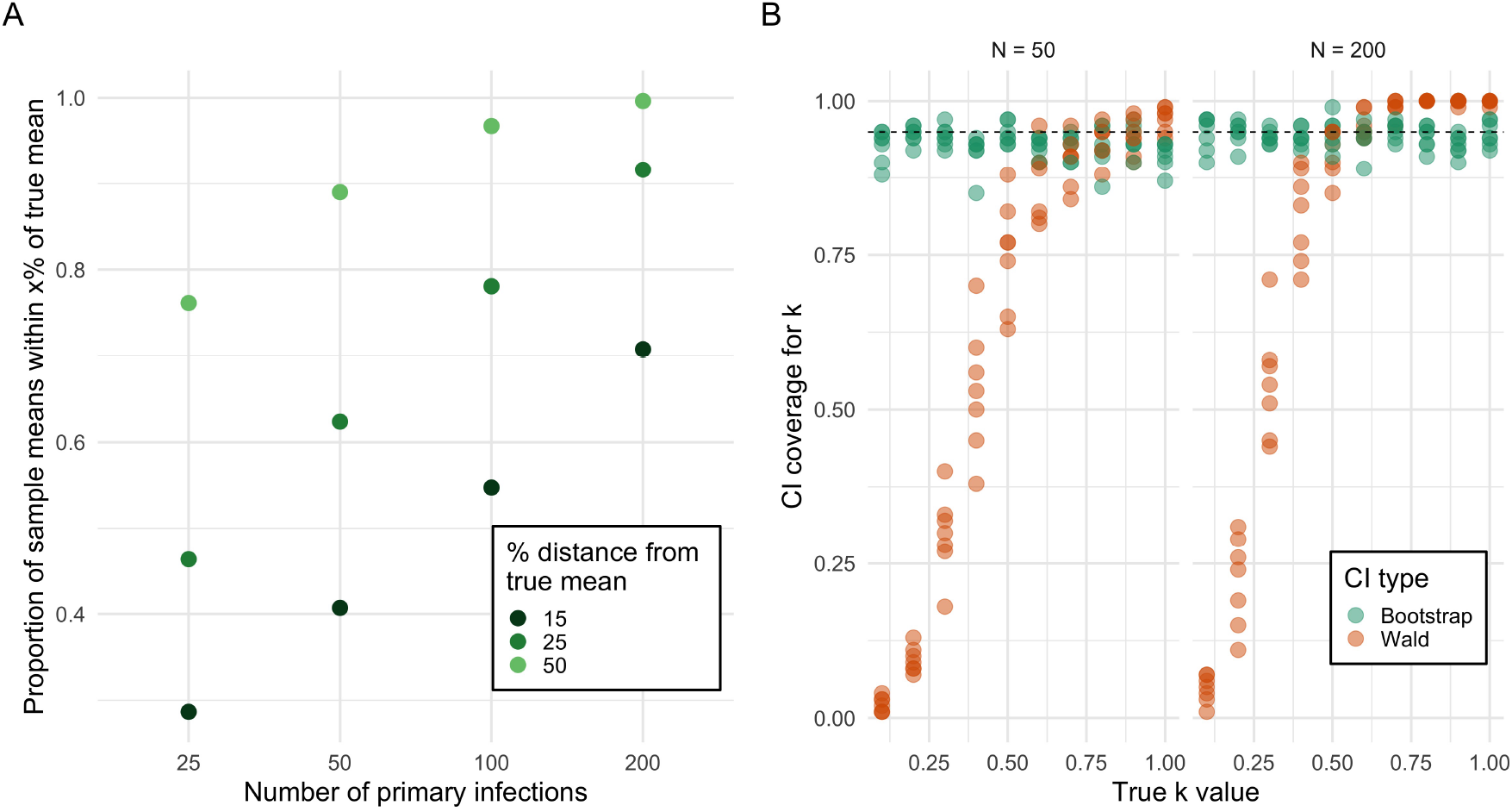
Point estimates and confidence intervals of *k* can be substantially inaccurate. a) Even assuming perfect random sampling, point estimates of *k* are noisy. With 100 observed primary infections (corresponding to 250-350 secondary infections), point estimates have only a 50% chance of being within 15% of the true mean. b) Confidence interval coverage guarantees depend strongly on the confidence interval method. The Wald interval does not match its coverage guarantees when the true *k* is small, even at large sample sizes. The non-parametric bootstrap method produces intervals that match the coverage guarantees regardless of sample size or true population *k* value. Dashed line indicates nominal 95% coverage.

We reanalyze publicly available contact tracing data from recent COVID-19 contact tracing studies ([19, 29, 30, 28]), and find that 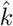 estimates are sub-stantially above the consensus range of 0.1-0.3 (Figure 4). There is substantial variation across the 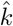 estimates, which vary in location, data collection practices, and local stage of the epidemic, among other differences. Despite this variation, the majority of the estimates are in the range 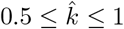 1. While the true bias in these estimates is unknowable, we also perform a minimal bias correction (Appendix Figure S5, Table S1), demonstrating that some of the most extreme estimates of *k* could overestimate superspreading. This bias correction is ad hoc and approximate – it should only be taken as an example of the potential magnitude of bias in the estimates. Even after the bias correction, the calculated range corresponds to substantial superspreading.

**Figure 4:**
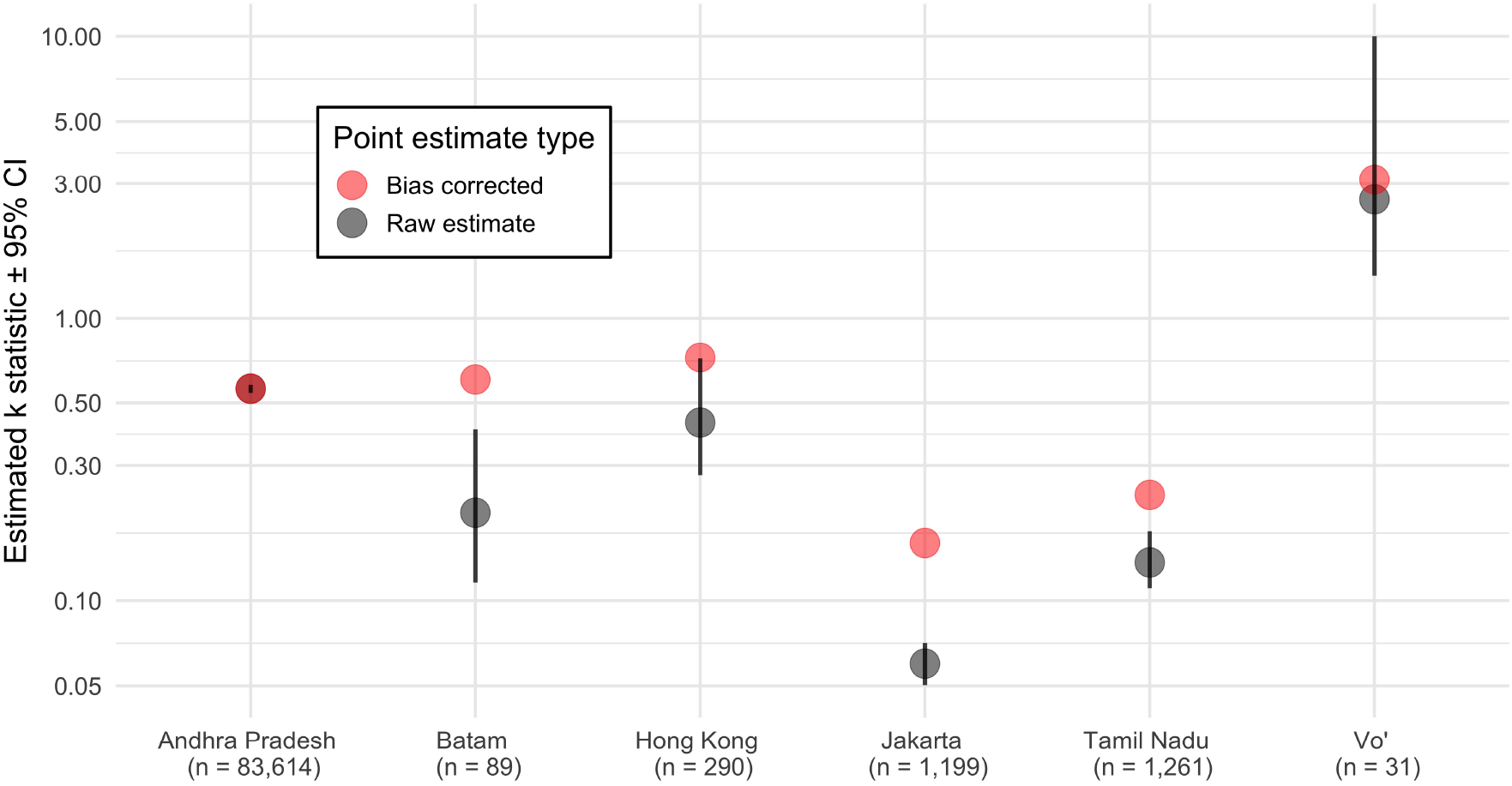
Reanalysis of contact tracing datasets shows 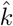 higher than conventional range of 0.1 − 0.3. Raw estimates (black) are based on maximum likelihood estimation with confidence intervals using a non-parametric bootstrap. Ad hoc bias corrected values are also shown (red). Data on Andhra Pradesh and Tamil Nadu are both from [28], on Jakarta and Batam are from [29], on Hong Kong are from [19], and on Vo’ are from [30]. The upper bound of the Vo’ confidence interval is truncated at 10 because for practical purposes *k* ≥ 10 produces a Poisson offspring distribution. The number of primary infections observed in contact tracing is listed below each study location.

## Discussion

The transmission of infectious agents within host populations is influenced by a number of sources of heterogeneity, including variation in behavior, immunology, physiology, and genetics. A consequence of such heterogeneity is disproportionate transmission dynamics in which a few hosts propagate pathogen efficiently, while the majority transmit inefficiently. In this work, we generate a mechanistic understanding of superspreading, uncovering that an extreme level of variation in underlying behavioral or biological processes is needed to achieve recent empirical estimates of superspreading potential. Such extreme levels of variation are not, however, supported by available behavioral or biological data. To explain this discrepancy, we consider how measurement biases may affect the quantification of superspreading potential from epidemiological data.

Our mechanistic approach shows that superspreading can be reproduced through the combination of heterogeneity in social contact and heterogeneity in transmissibility across the population. While the combination of these simple phenomena is important to produce superspreading, the process is largely governed by the amount of variation in social connectivity present in the population. The relationship between social contact and superspreading is remarkably robust; the relationship between transmissibility and superspreading is much more fragile. While variation in these factors is critical, superspreading (estimation of *k*) is largely robust to the transmissibility and degree means. However, as is well known, these means are key elements of epidemic spread, affecting *R*_0_ and the probability of an epidemic, among other important metrics.

Our results indicate that variation in transmissibility alone, even when extreme variation, may fail to accurately describe superspreading. When relying solely on variation in transmissibility, these descriptions are extraordinarily sensitive to the exact transmissibility parameterization and can easily produce unstable and unrealistic estimates (for example: *k >* 5). A population with variability in transmissibility but not behavior is analogous to a homogeneous-mixing compartmental model ([14]), and superspreading inference from these homogeneous-mixing models should be treated with caution – their attempts to reproduce superspreading may be overly idealized. Indeed, careful empirical modeling of SARS-CoV-2 superspreading has emphasized the importance of social contact to accurate description [31, 32, 33].

The robust qualitative relationship between *k* and the amount of social contact variability in the population suggests empirical estimates of *k* can provide information about the social contact network along which the pathogen spreads. Pathogens that exhibit little to no superspreading behavior theoretically correspond to a population with little degree heterogeneity (e.g. a Poisson degree distribution). A pathogen exhibiting minimal superspreading events (i.e.1 ≤ *k* ≤ 1.5) corresponds to a over-dispersed degree distribution, while one with substantial superspreading (0.5 *< k <* 1) corresponds to substantial degree variability. We argue that extreme superspreading (*k* ≤ 0.3) strongly supports a scale-free degree distribution.

Past work has suggested that highly variable degree distributions may describe the contact networks of sexually transmitted diseases, but may not be representative of respiratory disease contact networks [14]. Contact networks for respiratory transmission based on a contact definition of a face-to-face conversation of three or more words or physical contact are described by relatively homogeneous degree distributions [34, 35]. However, the ubiquity of superspreading behavior in the dynamics of respiratory-transmitted coronaviruses such as SARS-CoV-1, MERS, and SARS-CoV-2 ([2, 36, 19, 37, 24]), suggests that the behavior underlying transmission for these pathogens may display more population variability than can be explained by existing contact definitions. Such variability may result from complexity in contact definitions (e.g. varying duration of contact or cumulative contact periods) or multiple transmission modes (e.g. droplet and airborne) driving transmission. The strong relationship between population contact variability and superspreading points to unexplained mechanisms of transmission for coronaviruses. Future work to address this question is critical.

As first demonstrated in [6], there is a robust relationship between quantitative estimates of superspreading (the *k* statistic) and adherence to the 80/20 rule, a scenario in which 80% of all infections are caused by 20% of infected individuals. Our first-principles model is able to reproduce this relationship, demonstrating that adherence to the 80/20 rule implies quantification of *k* ≈ 0.3. These results imply that when datasets “satisfy” the 80/20 rule, they should likely produce estimates of *k* in the vicinity of 0.3, as occurs empirically in [15]. The opposite is also true – estimates of *k* above 0.3, while overdispersed, should not adhere to the 80/20 rule.

The theory of the negative binomial offspring distribution makes the practice of quantifying superspreading potential from a dataset quite feasible, but the method itself is noisy and subject to bias – results should be interpreted with care. There are inherent methodological challenges in estimating a variance parameter as it approaches an extremum; the negative binomial dispersion parameter has been the subject of particular attention because of its relatively flat likelihood, which makes accurate estimation and computation substantially more challenging [38, 39]. The combination of noisy point estimates, poor confidence interval coverage, and non-random sampling can produce overconfident underestimates of the *k* dispersion parameter. For this reason, sources of bias and the full range of the bootstrap confidence interval must be considered with care. Empirical estimation of *k* also suffers from inherent selection bias, with datasets chosen for analysis because they demonstrate superspreading events. This process, while well-intentioned, induces statistical flaws, especially for small or only partially observed samples. The combination of several superspreading events into one larger dataset only compounds the issue (Appendix Figure S6). While ad hoc bias correction can help mitigate this source of bias, the best way to produce accurate estimates is to use large, rigorous samples as in [28].

In our empirical reanalysis, we find *k* in the range of 0.5 to 1 for COVID-19, which indicates substantial superspreading, and agrees with some thorough empirical estimates [31]. However, there is notable variability in estimates of *k*, both across settings and within studies across multiple sites. This variation is to be expected; at the time of observation, the municipality of Vo’ had a large portion of the population already infected and implemented strict social distancing procedures, likely altering estimates of *k* [30]. *On the other hand, estimates of k* in Indonesia are much lower, but should be interpreted with extreme caution. While the Indonesian study was conducted in the middle of uncontrolled epidemic growth, the naive *R*_0_ estimates associated with the *k* estimation procedure were below 1 and tests to confirm cases were scarce, especially in Jakarta, which both point to substantial under-reporting and potential biases in data collection [29]. In addition, both [28] and [29] exhibit within-study heterogeneity in *k* estimates across geographic region and while these estimates are certainly influenced by variation in local public heath capacity, this heterogeneity also suggests that local differences in population structure could alter superspreading rates (as implied by [40] and [31]). Similarly, there were substantial differences in numbers of tests and test positivity rate between Tamil Nadu and Andhra Pradesh, potentially leading to biases in linelist inclusion [28]. Differences in factors such as public health interventions (i.e. NPIs, social distancing), local epidemic scale and spread, and linelist inclusion criteria, among others, could all contribute to the substantial unexplained differences between empirical superspreading estimates. These limitations point to the need for public availability of large-scale contact tracing datasets and a more thorough examination of biases in *k* estimation.

Our results suggest that public health interventions meant to decrease the rate of superspreading events (e.g. social distancing, bar closures) may not materially alter measurements of superspreading (i.e. *k*) even though they are effective at decreasing infection rates. These interventions are undoubtedly important techniques in the public health arsenal, reducing rates of spread and total disease burden, and should be diligently applied and followed. However, a superspreading pathogen (*k <* 1) is dependent on substantial contact heterogeneity and even large reductions in contact heterogeneity are unlikely to decrease *k* by more than 0.1-0.3 when the initial *k* is less than 1; rapid decrease of *k* only occurs at smaller values of contact heterogeneity which do not produce much superspreading. For this reason, interventions such as social distancing may not meaningfully decrease measurements of *k* in superspreading diseases, even though they provide substantial benefit through reduction in *R*_0_. Interventions to decrease transmissibility (e.g. mask wearing) may also have limited impact on superspreading quantification due to the insensitivity of *k* to transmissibility. However, even though these interventions may not decrease *k*, they may decrease superspreading – because *R*_0_ and *k* are dependent on each other, descriptors of the offspring distribution, a reduction in *R*_0_ even without a reduction in *k* corresponds to smaller superspreading events. Indeed, superspreading pathogens are sensitive to mitigation strategies that preferentially prevent superspreading events, with reductions in behavioral variability providing superior reduction in *R*_0_ [41, 42, 27]. Research on the impact of superspreading diseases to public health interventions is still an emerging field, with substantial uncertainty remaining in how to best implement interventions.

These results, based on theory and mathematical modeling, build upon on an extensive and rapidly growing literature on epidemic spread in complex systems – an area of immediate public health concern. A strong understanding of heterogeneity in disease transmission dynamics is not only vital to the public health response to SARS-CoV-2, but also critical to the response to future epidemics. Disease spread is influenced by many different sources of real-world heterogeneity; understanding and accurately quantifying disease dynamics is impossible without robust development of epidemic theory.

## Methods

### Epidemic network model

To model disease transmission in a population with behavioral and/or biological variability, we consider a network-based epidemiological model. In the network, nodes represent individuals, and edges between nodes represent potential disease-causing interaction (e.g. shaking hands, a conversation in close proximity).

We represent the population as a random network with a specified degree distribution. Most of our analysis involves (static) random networks with a negative binomial degree distribution, parameterized with a mean and variance parameter which can be tuned to a specified index of dispersion (i.e. variance to mean ratio). To understand edge cases, we also consider regular random networks (in which all nodes have the same degree) and scale-free random networks (in which the degree distribution is a power law). We generate random graphs of size *N* = 5000 in Python using the package “networkx”. All degree distributions are shifted such that the minimum degree is 1, and all networks are constrained to be simple (no self loops).

We simulate disease transmission in a population based on the percolation Susceptible-Infectious-Recovered (SIR) model. Disease spreads on each network edge with a transmissibility, *T*, which is a probability of transmission between an infected and susceptible individuals [43]. We seed one index patient and allow the infection to propagate until there are no infected individuals remaining. We classify an outbreak as an epidemic if a minimum of 5% of the population is infected. We report the outcomes as an average across 1000 Monte Carlo simulations. We use the simpler SIR model over the more complicated Susceptible-Exposed-Infectious-Recovered structure because the Exposed class alters the time to infectiousness for an individual but does not change the number of infections per infector, which is the metric used for estimation of *R*_0_ and *k*. For each set of Monte Carlo simulations, we record the offspring distribution at the second generation, and fit a negative binomial distribution to the aggregated offspring distribution of the epidemics using maximum likelihood estimation to get the mean (*R*_0_) and the dispersion statistic (*k*) of the distribution [38]. We also calculate the proportion of total infections due to the 20% of individuals with the most realized infections in the offspring distribution and the per-introduction probability of an epidemic.

We consider two sets of experiments to understand the role of behavioral vs biological factors in generating superspreading. First, we alternatively assume variability in either behavior (degree) or biology (transmissibility). To achieve this, we either assume negative binomial distributions for degree with a fixed mean of 8 and a varying index of dispersion, and a constant transmissibility with *E*[*R*_0_] = 3. Alternatively, we assume a homogeneous degree of 8, and a heterogeneous transmissibility parameterized by scaled negative binomial distributions so that the support is in [0, 1]. Second, we simultaneously assume variability in degree and transmissibility, while allowing *R*_0_ to vary within *R*_0_ ∈ [1, 5].

### Maximum likelihood estimation and confidence intervals for *k*

To examine the accuracy of maximum likelihood point estimates and confidence interval coverage for the *k* statistic, we consider a negative binomial offspring distribution across a range relevant biological scenarios and examine the accuracy of the fit and associated error. For every combination of negative binomial dispersion statistic, *k* ∈ {.1, .2, …, .9, 1}, negative binomial mean, *R*_0_ ∈{2, 2.5, …, 4.5, 5}, and sample size, *N* ∈ {25, 50, 75, 100, 200}, we draw negative binomial samples, use maximum likelihood to estimate the parameters, 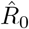 and 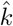, and generate the associated Wald and naive non-parametric bootstrap confidence intervals for 100 Monte Carlo simulations. We follow the method employed in [38], calculating the likelihood and associated statistics about *k* on the *k*^−1^ scale.

We also explore the effect of non-random sampling and imperfect observation on estimation of *k*. We simulate the impact of sample *selection* (i.e. the practice of only choosing datasets exhibiting superspreading to estimate *k*) in the presence of imperfect contact tracing. We generate samples from a negative binomial offspring distribution with *R*_0_ = 3 and *k* ∈ {0.5, …, 1} with a binomial observation process with probability of observation *p*_*obs*_ ∈ {0.15, 0.25, 0.5, 0.75}. The resulting generative structure is:

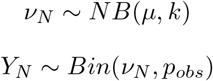

We use this structure to compute the average bias caused by selection (i.e. non-random selection of samples) if the true offspring distribution *k* were in the range [0.5, 1], corresponding to substantial but not extreme superspreading. This average effect is not evidence for bias; rather, it is an estimation of the direction and magnitude of this potential bias if it were occurring.

### Analysis of contact tracing datasets

Using publicly released COVID contact tracing datasets [19, 29, 30, 28], we calculate the dispersion parameter *k* using maximum likelihood estimation and associated 95% confidence intervals using the non-parametric bootstrap. We apply the ad hoc bias correction for *p*_*obs*_ = 0.15 from the previous section to the estimates. We process and analyze the datasets in R using the package ‘fitdistrplus’.

## Data Availability

All simulation code, analysis code, and data are made available at https://github.com/zsusswein/COVID_superspreading.

## Acknowledgements

We acknowledge valuable technical feedback from Sam Scarpino, Greg Albery, Eva Rest, and Jack Townsend. Research reported in this publication was supported by the National Institute Of General Medical Sciences of the National Institutes of Health under Award Number R01GM123007. The content is solely the responsibility of the authors and does not necessarily represent the official views of the National Institutes of Health.

## Supplementary Information

**Figure S1:**
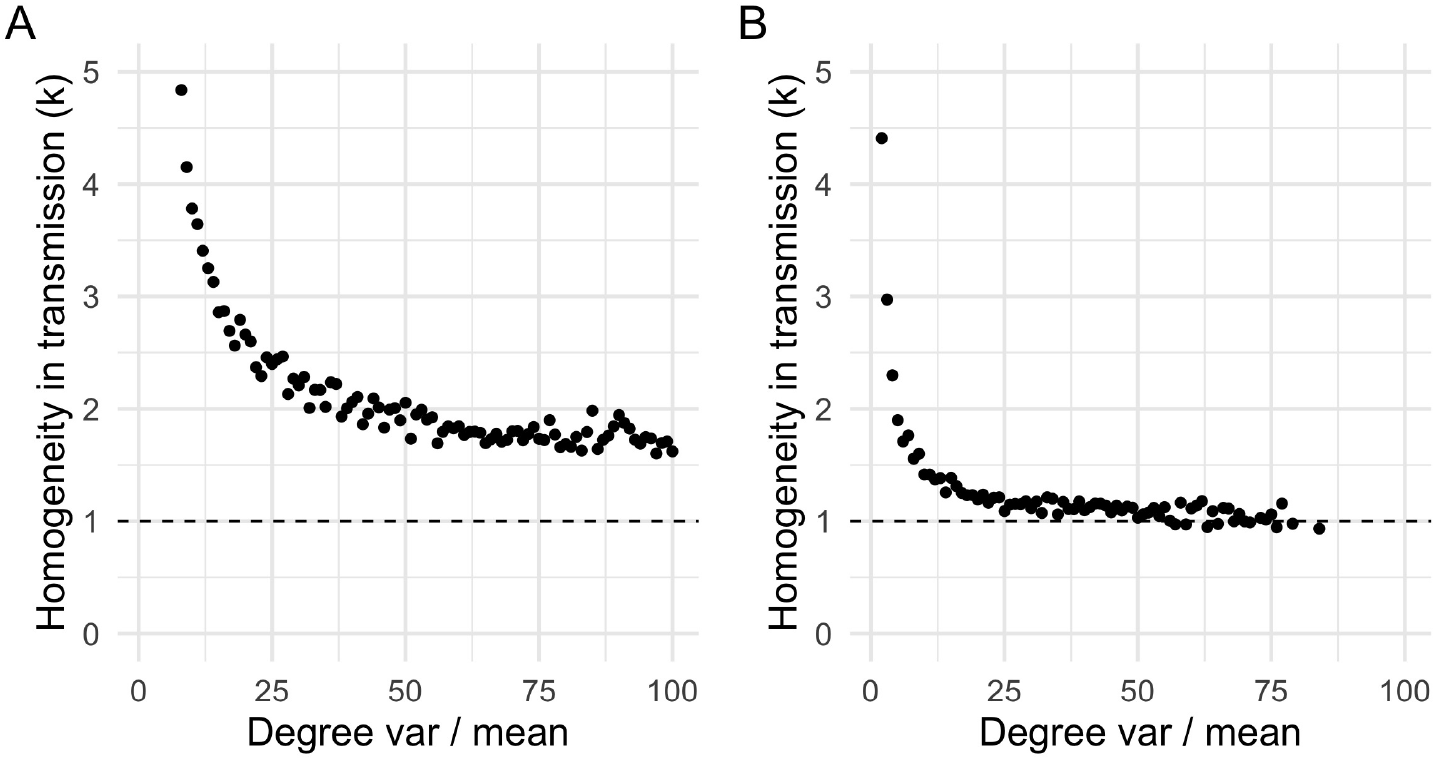
The mean of the degree distribution alters the convergence rate to the asymptote but does not change the minimum *k* statistic in the univariate case. a) A degree distribution with mean of 16 has *k* decrease more slowly with behavior heterogeneity than for mean degree of 8. b) A degree distribution with mean of 4 has *k* decrease more quickly than a degree distribution with mean of 8. Despite the more rapid decrease the *k* statistic does not decrease below the asymptote of *k* = 1.

**Figure S2:**
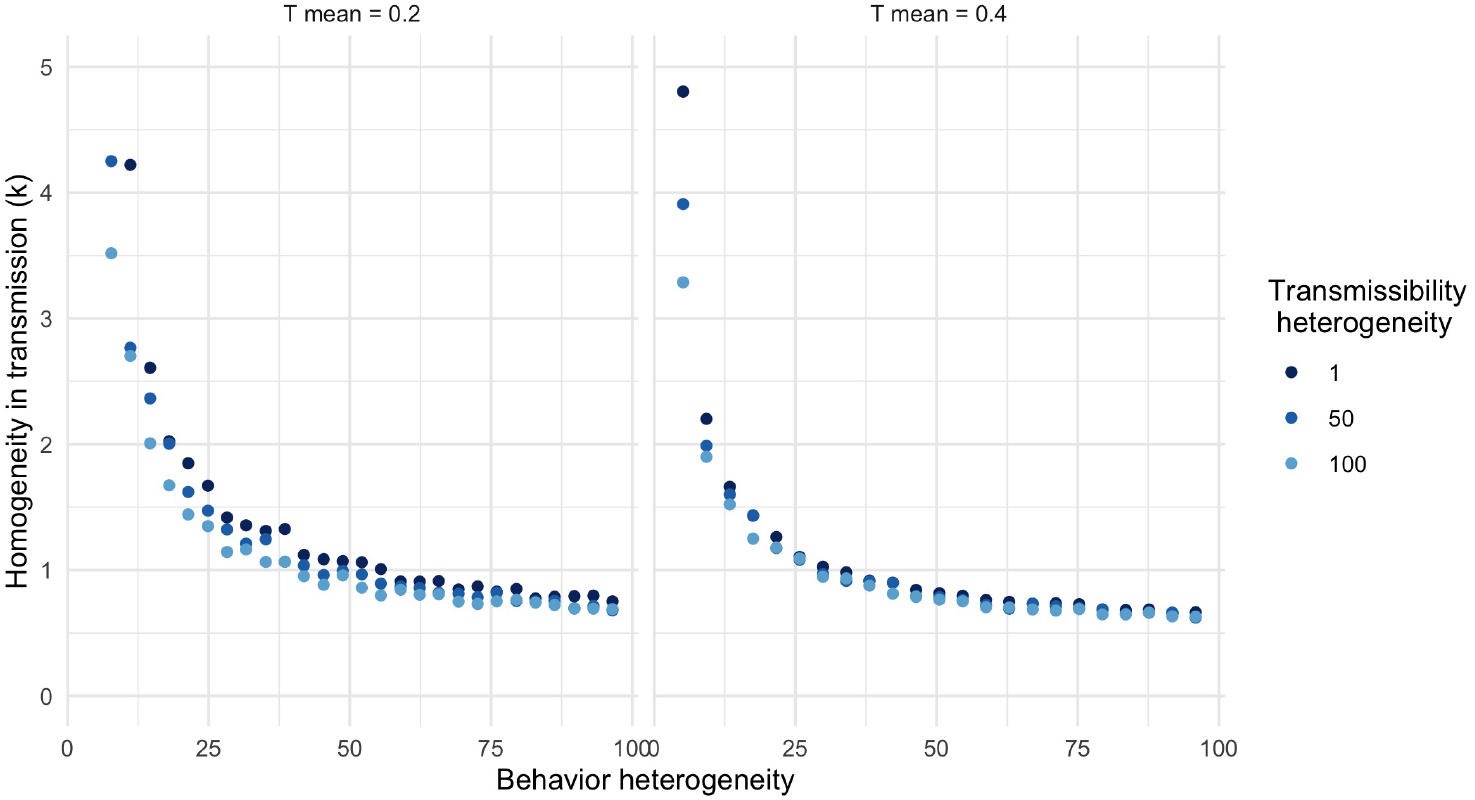
The k statistic is only weakly affected by the transmissibility mean. Increases in the mean transmission probability lead to faster convergence to the asymptotic regime (*k* ≈ 0.5), but do not lead to *k* below 0.5.

**Figure S3:**
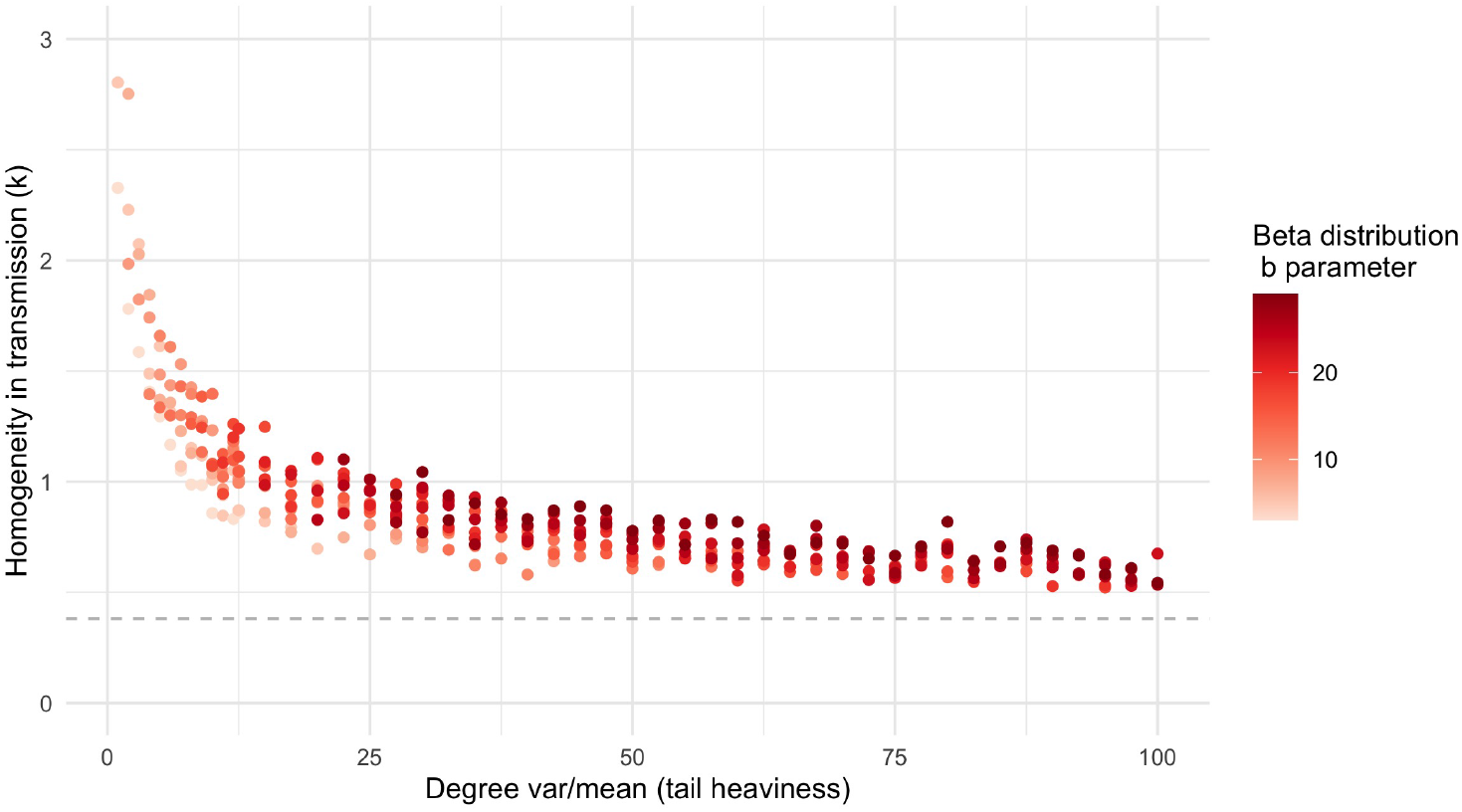
Superspreading estimates are invariant to the distributional form of transmissibility. Using beta distributions with shape parameter a = 1 and varying shape parameter b, *k* declines exponentially with behavior heterogeneity. Superspreading dynamics are governed by asymptote, *k* = 0.5, as when transmissibility represented by scaled negative binomial distributions. A scale-free degree distribution (grey dashed line) produces *k* ≈ 0.3.

**Figure S4:**
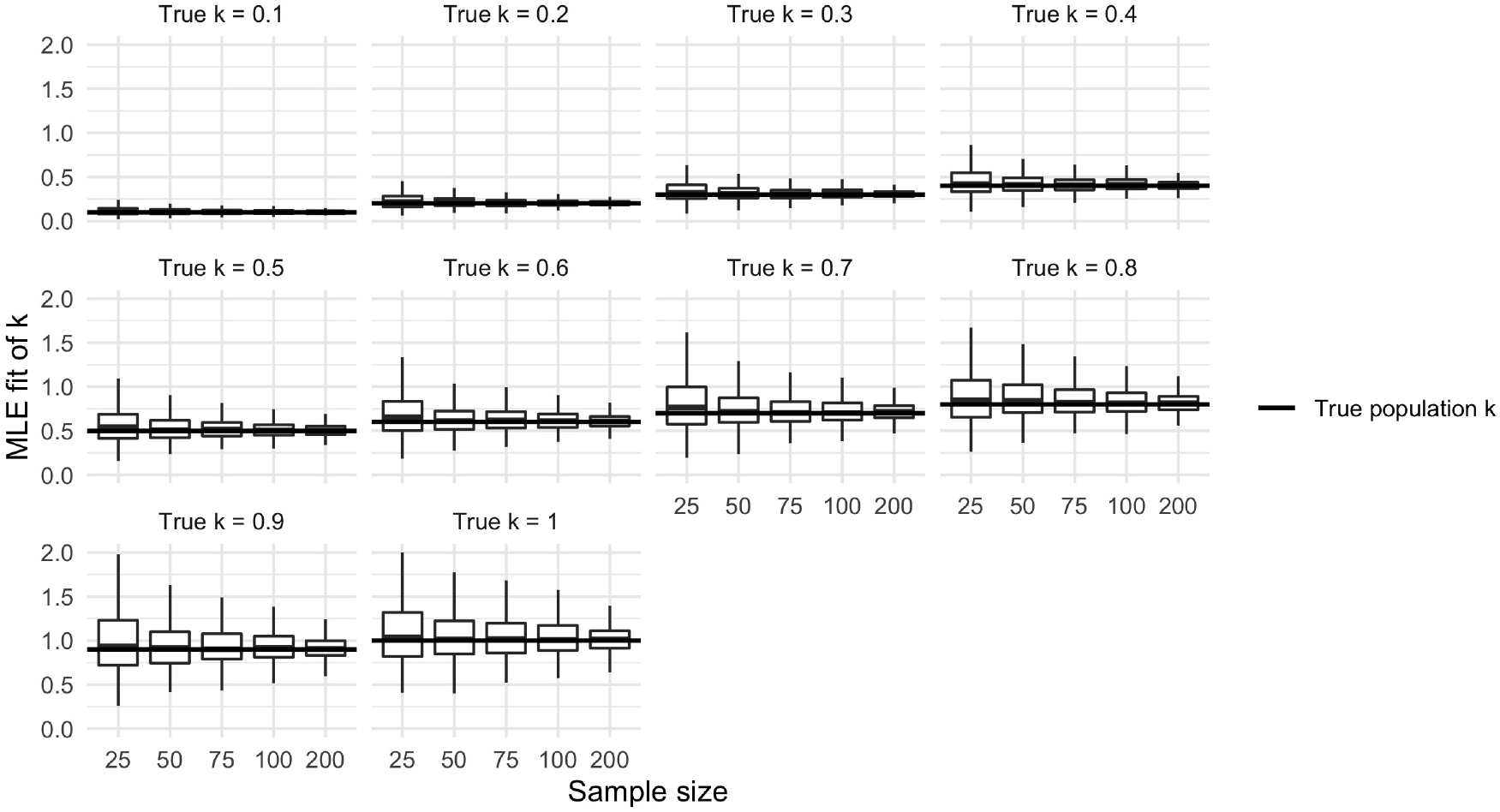
Estimates of *k* drawn from random samples are noisy and symmetric. The variability of *k* estimates increases with true value of *k*. When the true *k* is small (*k* ≤ 0.5), estimates are tightly concentrated around the true value, even at smaller sample sizes. When the true *k* is moderately larger (0.5 *< k* ≤ 1), the estimates of *k* are much more variable and can produce smaller values of *k* through sampling variability alone.

**Figure S5:**
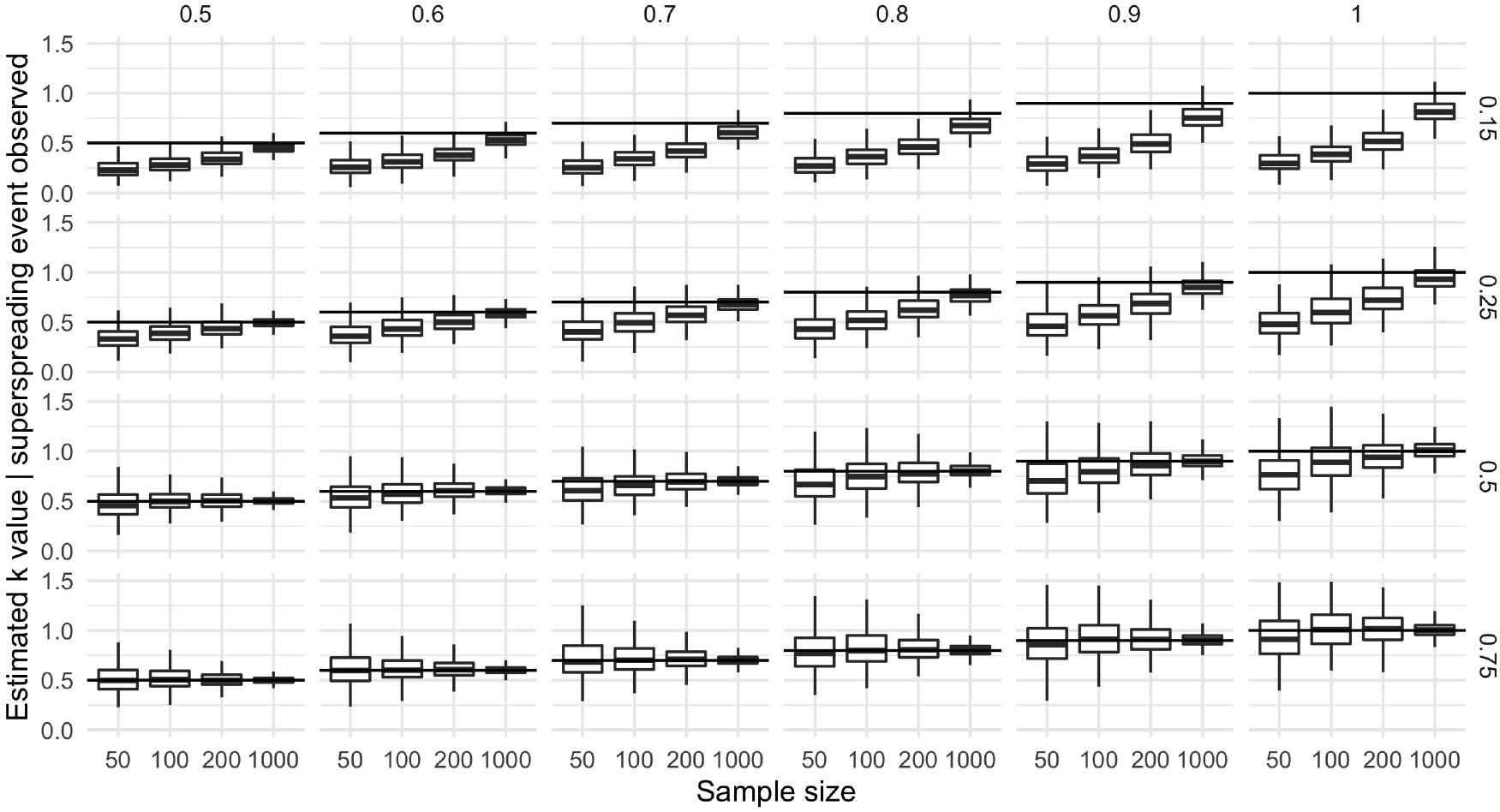
Sampling bias can lead to substantial underestimates of *k*, compounded by imperfect observation. When cases are under-reported, estimates of superspreading using only datasets that include a superspreading event (greater than 8 infections [6]) are substantially biased to overestimate superspreading. This bias is magnified by the proportion of cases observed (right axis) and the true *k* value (top axis). This bias persists even at larger sample sizes.

**Table S1:** The mean bias from sample selection can lead to substantial underestimates of *k*. As described in the methods, samples from a negative binomial offspring distribution with *k* ∈ [0.5, 1] produce biased estimates if only samples with superspreading events are analyzed. This bias is magnified by small sample sizes and under-reporting.

| **N** | **Secondary case detection probability** | **Bias** |
| --- | --- | --- |
| 50 | 0.15 | 0.47 |
| 50 | 0.25 | 0.32 |
| 50 | 0.50 | 0.09 |
| 50 | 0.75 | -0.01 |
| 100 | 0.15 | 0.40 |
| 100 | 0.25 | 0.23 |
| 100 | 0.50 | 0.04 |
| 100 | 0.75 | -0.03 |
| 200 | 0.15 | 0.30 |
| 200 | 0.25 | 0.15 |
| 200 | 0.50 | 0.00 |
| 200 | 0.75 | -0.02 |
| 1000 | 0.15 | 0.10 |
| 1000 | 0.25 | 0.03 |
| 1000 | 0.50 | -0.01 |
| 1000 | 0.75 | 0.00 |

**Figure S6:**
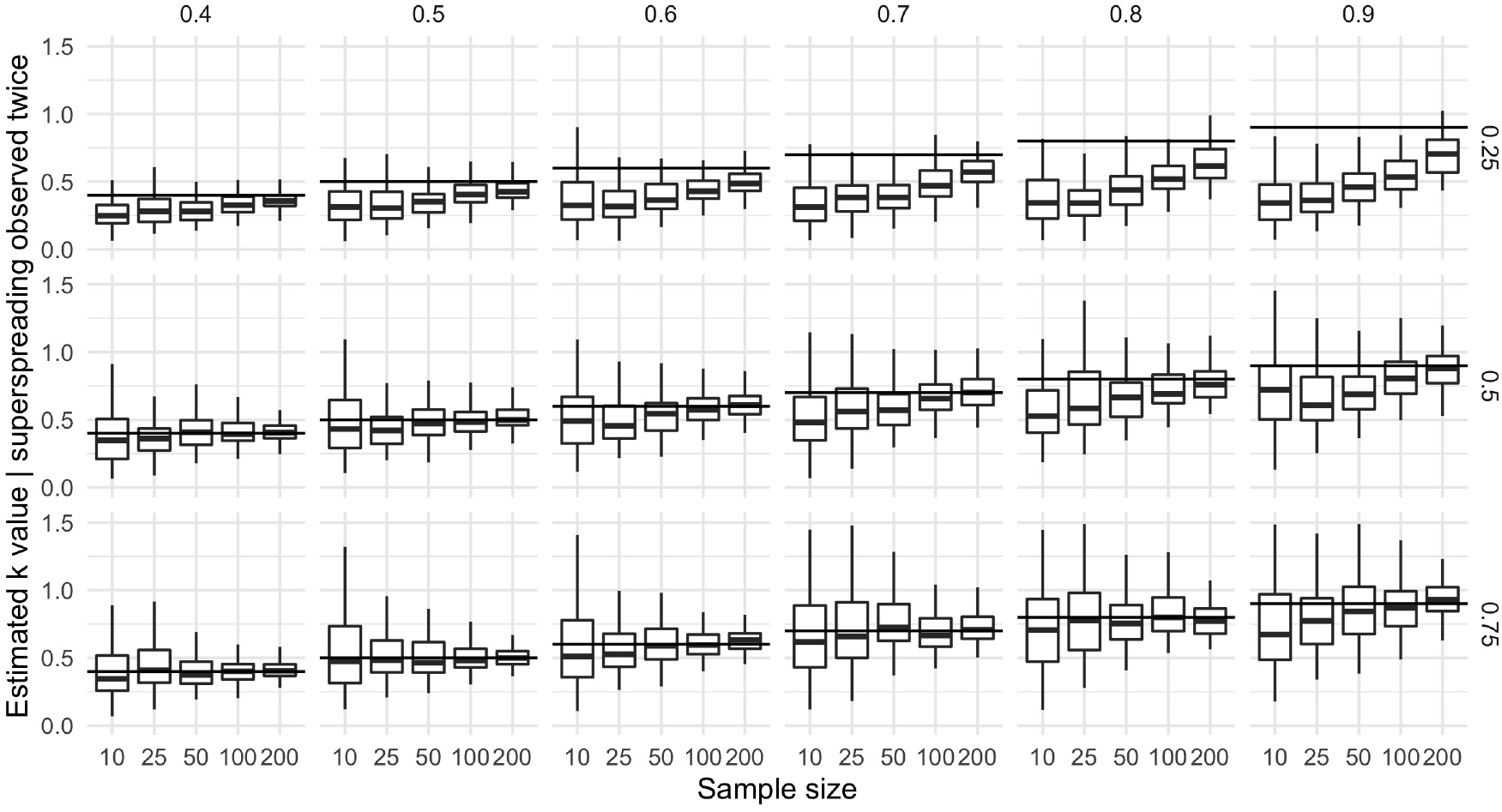
Sampling bias is magnified by the combination of two observed super-spreading events. The combination of two selected datasets does not mitigate the bias induced by the selection procedure. As in the case with one observed event, this bias is magnified at small sample sizes, at larger true vales of *k* (top axis), and at lower proportions of cases observed by contact tracing (right axis)

